# EFFICACY AND SAFETY OF NICORANDIL IN ACUTE CORONARY SYNDROME: A META-ANALYSIS

**DOI:** 10.1101/2023.04.04.23288145

**Authors:** Avichal Dani, Shubh Raithatha, Dev Desai

## Abstract

**Background:** Nicorandil acts as a potassium channel opener, however its cardio protective benefit is still uncertain. This meta-analysis was conducted with the objective of evaluating the efficacy of nicorandil in improving cardiovascular outcomes in acute coronary syndrome.

**Main Body:** A total of 24 RCTs with 1640 patients in the nicorandil group and 1592 patients in the control group were identified following PRISMA guidelines till November 2019 and were matched for inclusion and exclusion criteria. The following search strings and MESH terms were used: “nicorandil”, “ACS”, “MACE”. Following this, nicorandil was evaluated for its efficacy and safety outcomes. RevMan 5.3 was used for appropriate statistical tests. Fixed and Random Effect Model Test were used and p<0.05 was considered statistically significant.

**Results:** Administration of nicorandil was found to be associated with a significant decrease in MACE (RR = 0.686, 95% CI = 0.509-0.925, p=0.013), no-reflow phenomenon (RR =0.395, 95% CI = 0.266-0.588, p<0.001) and worsening of HF (RR =0.441, 95% CI = 0.221-0.882, p=0.021). It was also associated with significant improvement in LVEF (SMD= 0.637, 95% CI= 0.0972 to 1.177, p=0.021) and significant lowering of cTFC (SMD= -0.216, 95% CI= -0.428 to -0.0041, p=0.046)

**Conclusions:** Nicorandil does indeed exert a cardio protective effect by improving cardiovascular outcomes. There is a significant decrease in occurrences of MACE and worsening of HF. There is also significant improvement in LVEF.

## Background

Acute Coronary Syndrome is a medical emergency and if not treated promptly can lead to Death of the patient. If mortality prevented, Myocardial injury, and Myocardial Death is the most important and grave complication of acute coronary syndrome and can precipitate another episode of ACS or can lead to rupture of myocardium and other array of problems.

Nicorandil is a medication that can be used to treat Acute Coronary Syndrome (ACS). Nicorandil is an ATP-sensitive K+ channel opener with antiarrhythmic and cardiac protective properties, including lowering micro-coronary artery flow resistance via relaxing blood vessel smooth muscles and preconditioning. (1) (2) nicotinamide nitrate, and has been shown to decrease infarct size and incidence of arrhythmias after coronary artery ligation and reperfusion in experimental animals.(3) Nicorandil is hypothesised to inhibit calcium input into myocytes and restrict membrane depolarization, hence reducing ATP consumption during ischemia, through its (ATP)- sensitive K+ channel opening action. (4)

Reperfusion of cardiac myocytes after the event has occurred and the patient has been saved is another hallmark of Acute Coronary Syndrome. Reperfusion damage is a major concern that can result in the patient’s mortality. As blood is reintroduced to the injured myocardial, locally produced toxins from the damaged or dead myocytes enter the bloodstream, causing harm to the renal and hepatobiliary systems.(5) (6)

Myocardial injury commonly occurs during percutaneous coronary intervention (PCI) and can be easily monitored by measuring myocardial enzymes. (4) it is a lifesaving procedure. (7) it can be seen with reduces the incidence of minor cardiac marker elevation after coronary stenting (4) PCI along with administration of Intravenous Nicorandil seems to be vastly beneficial. (3) (8) (2)

There are paper suggesting that Nicorandil improves the cardiac function and clinical outcome in a case of acute myocardial infarction (3) the same results can be seen with monitoring cardiac enzymes too (9)

Nicorandil is a widely used drug, yet its exact benefits are not known yet. Most importantly, its effect on major advanced cardiac events and left ventricular ejection fraction. It can be seen that as it is used in emergency, it is beneficial to reduce mortality but it is still a matter of question about its benefits on a long term period.

## Main Body

### Methodology

#### Data source

A computerized search was undertaken in PubMed, Cochrane Library, Google Scholar, and the Controlled Trials Meta Register. “Nicorandil”, “Acute Coronary Syndrome”, “MACE” and “LVEF: were text keywords. Additional studies were found by manually searching the reference lists of pertinent retrieved publications. The only language allowed was English. Human investigations were the only ones that yielded results.

#### Eligibility Criteria

The researchers looked studies that compared Nicorandil vs Control in treatment of acute Coronary Syndrome. Along with Abstracts, letters, comments, editorials, expert opinions, reviews without original data, and case reports, Research papers were also excluded from the analysis if (10) it was impossible to extract the appropriate data from the published articles; [2] there was significant overlap between authors, institutes, or patients in the published literatures; [3] the measured outcomes were not clearly presented in the literatures; [4] the measured outcomes were not clearly presented in the literatures; and [5] articles were written in non-English.

#### Study Identification

All titles and abstracts discovered by the search approach were reviewed by the author. Relevant complete publications were collected for detailed evaluation; two non-author independent reviewers separately assessed them for eligibility criteria.

#### Data extraction

Each qualified manuscript was reviewed by two independent reviewers separately. The number of patients, their age, gender, intervention type, outcome was all collected from each manuscript. Conflicts were addressed by further discussion or consultation with the author and a third party. The modified Jadad score was used to measure the study’s quality.

#### Outcome Measures

Outcomes like MACE, no-reflow phenomenon (NRP), LVEF(Left Ventricular Ejection Fraction), cTFC(corrected TIMI Frame Count) and Worsening heart failure were considered.

#### Statistical analysis

All of the information was gathered and placed into software for analysis. To analyze important clinical outcomes, fixed- or random-effects models were used to generate mean difference, standardized mean difference (SMD), odds ratios, and relative risk (RR) with 95 percent confidence intervals (CIs). Statistical heterogeneity was measured with the χ2; P < 0.100 was considered as a representation of significant difference. I2 greater than or equal to 50% indicated the presence of heterogeneity.

## Results

Nicorandil Administration was found to be associated with a significant decrease in MACE (RR = 0.686, 95% CI = 0.509-0.925, p=0.013), which can be supported by (3) (11) (8) (12), whereas (13) (14) strongly suggest otherwise. Nicorandil was also found to be associated with a decrease in worsening of HF (RR =0.441, 95% CI = 0.221-0.882, p=0.021) similar to (8) (15) but (16) is strongly against this concept. It was also associated with significant improvement in LVEF (SMD= 0.637, 95% CI= 0.0972 to 1.177, p=0.021) as seen with (3) but (9) says it worsens LVEF. Nicorandil was associated with decrease in no-reflow phenomenon (RR =0.395, 95% CI = 0.266-0.588, p<0.001) as seen in papers like (5) (17) (11) Nicorandil was found to be responsible for significantly lowering of cTFC (SMD= -0.216, 95% CI= -0.428 to -0.0041, p=0.046) similar to (3) (5)

## Discussion

Nicorandil is already one of the most important drug while battling acute coronary syndrome and the efficacy of the drug has always saved lives of many patients. The results that can be seen here are only a proof of that. a proof of how good Nicorandil is not only for short term life saving role but at a long term basis too.

As it can be seen in Figure 1, Nicorandil seems to be sufficient enough to not only save the life of the patient but is similarly good in preventing MACE so that patient can lead the life without again combating any major advanced cardiac event. This result can be supported by a number of papers like (18) (5) (12) (11) (8) and many more. We only found 3 papers who says that Nicorandil is actually a reason behind increase amount of MACE as they found it (16) (14) (13)

**Figure 1:**
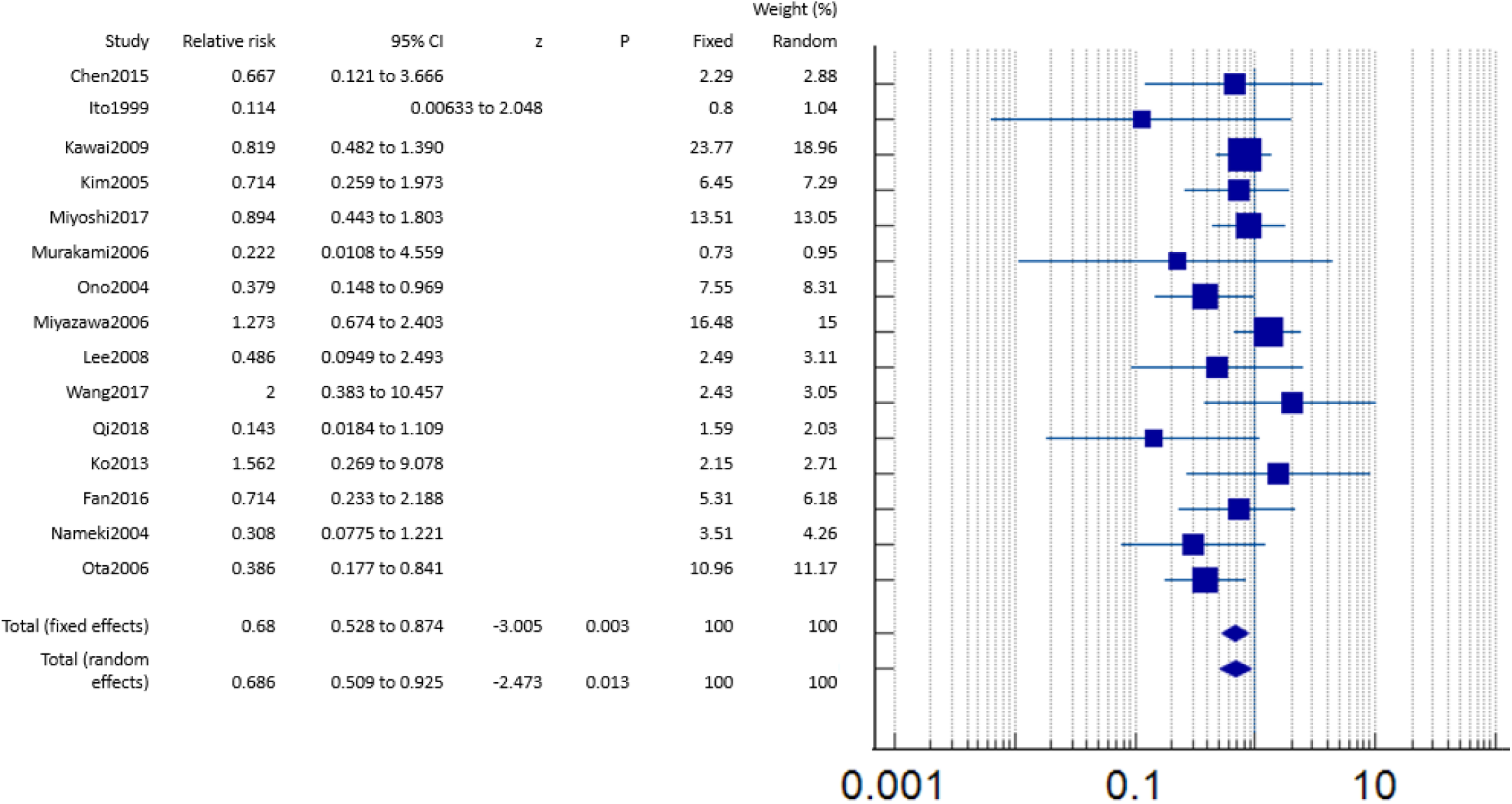
MACF Comparison 1: - Major Advanced Cardiac Event (MACE)

Figure 2 helps us understand what happens to the already existing heart failure condition when Nicorandil is administered. It is seen with many papers like (8) (3) (15) (14) (11) that Nicorandil should be a drug given to prevent the worsening of heart failure. Only 2 papers contrasted to it (16) (18), but the results of these papers very strongly shows that Nicorandil actually worsens heart failure. Altogether, Nicornadil seems to be helping in preventing the worsening of heart failure.

**Figure 2:**
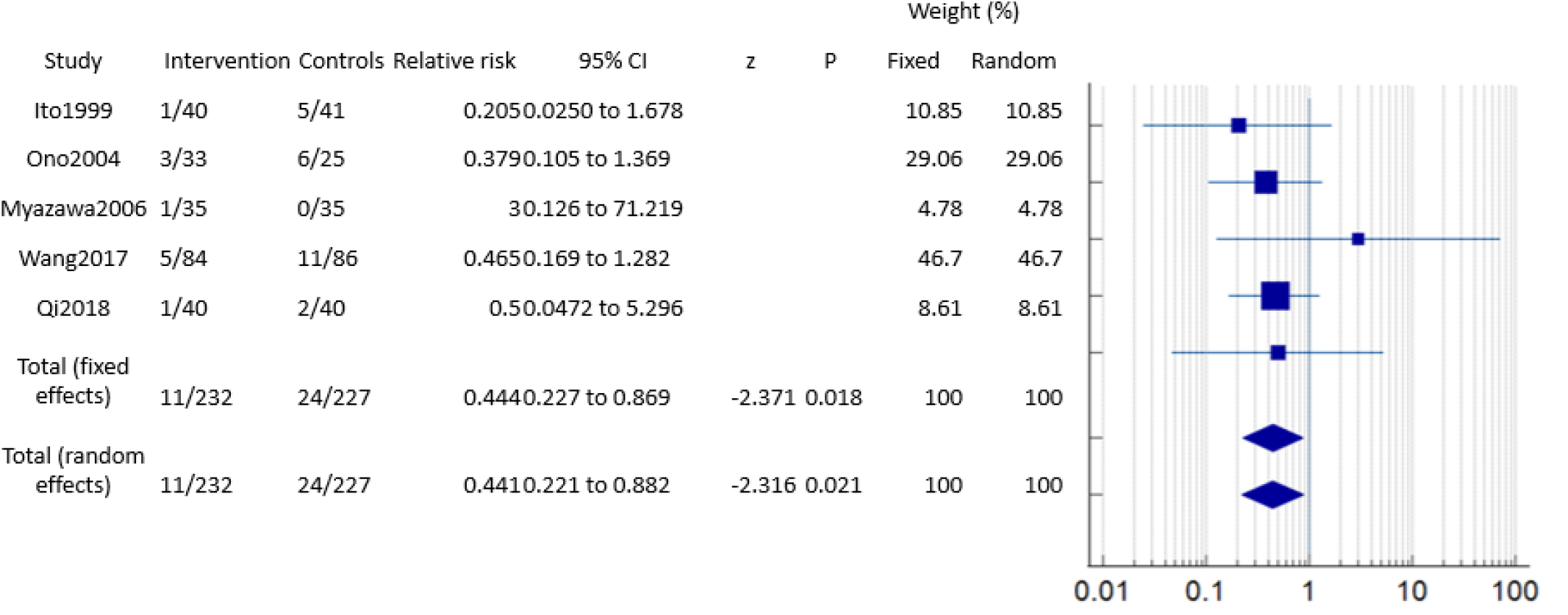
WHF Comparison 2 :- Worsening Heart Failure (WHF)

Overall, a positive impact on LVEF is seen with administration of Nicorandil as per Figure 3. An increase in LVEF is shown by papers like (3) (12) (16) (1) and more. Still, there were some papers like (19) showed that Nicorandil decreases the left Ventricular Ejection Fraction.

**Figure 3:**
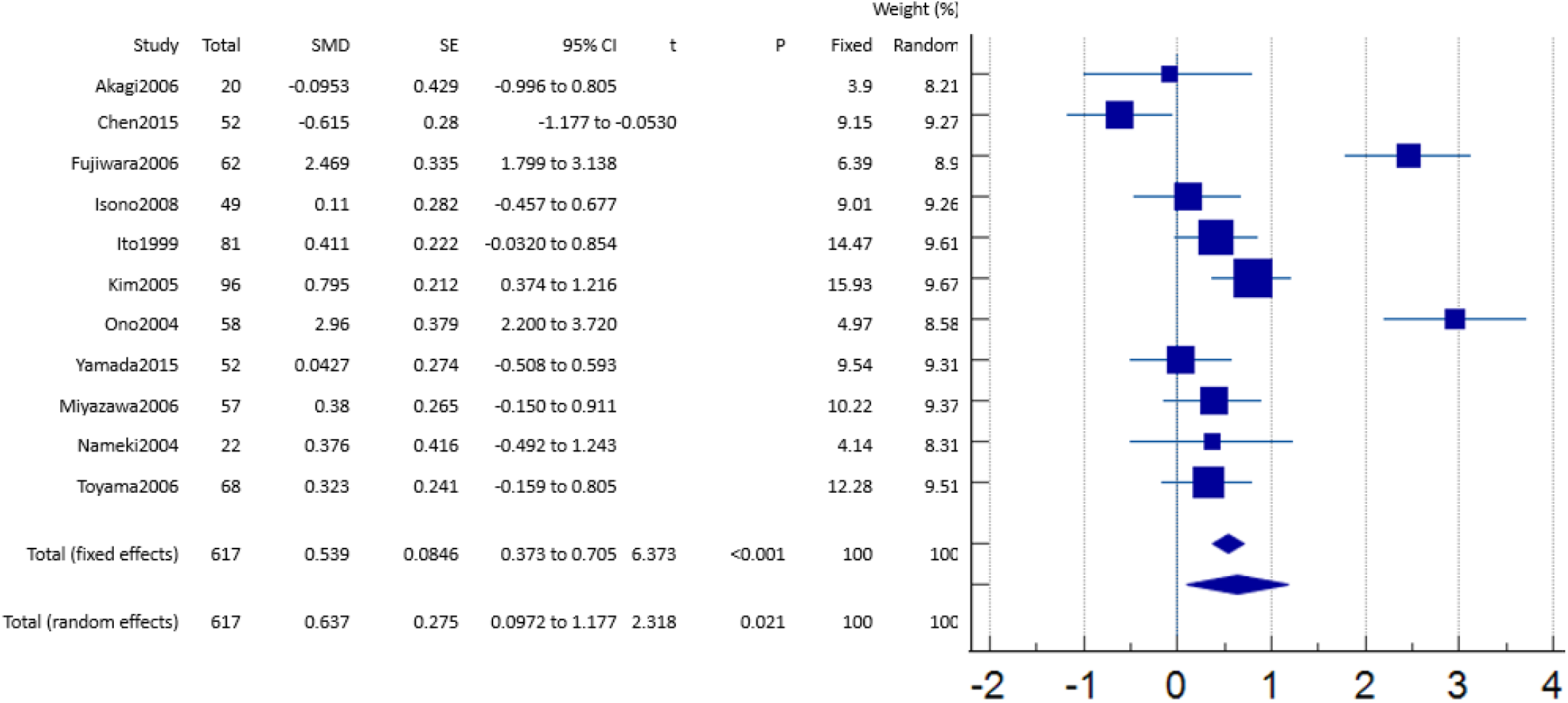
LVEF Comparison 3: - Left Ventricular Ejection Fraction

No-reflow phenomenon seems to be reducing with administration of Nicorandil as seen in figure 4. Papers like (5) (14) (17) (3) and others strongly suggest the protective role of Nicorandil. Almost all papers found show that Nicorandil decreases the phenomenon.

**Figure 4:**
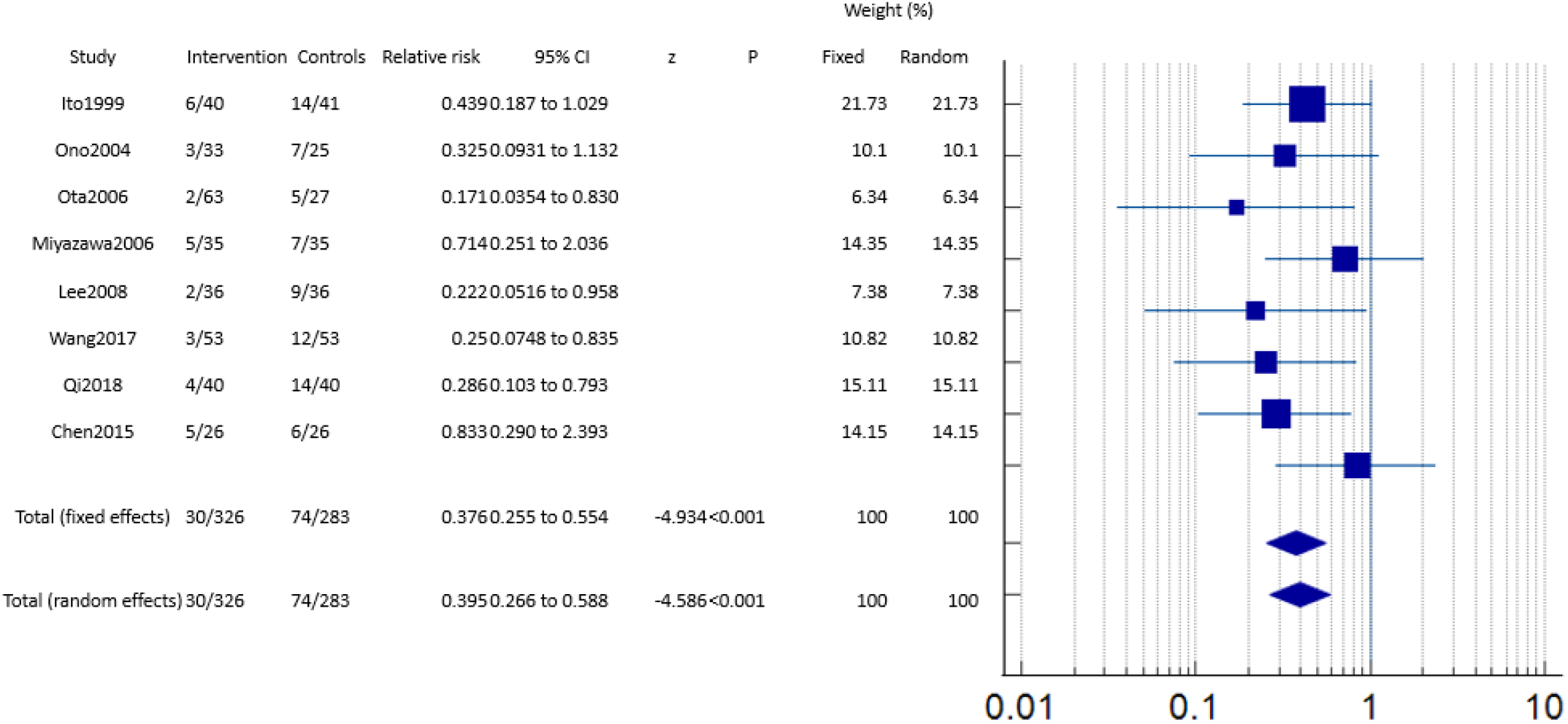
NRP Comparison 4: - No-Reflow Phenomenon (NRP)

Figure 5 shows a significant reduction in cTFC, and almost all papers like (3) (14) (5) support this result.

**Figure 5:**
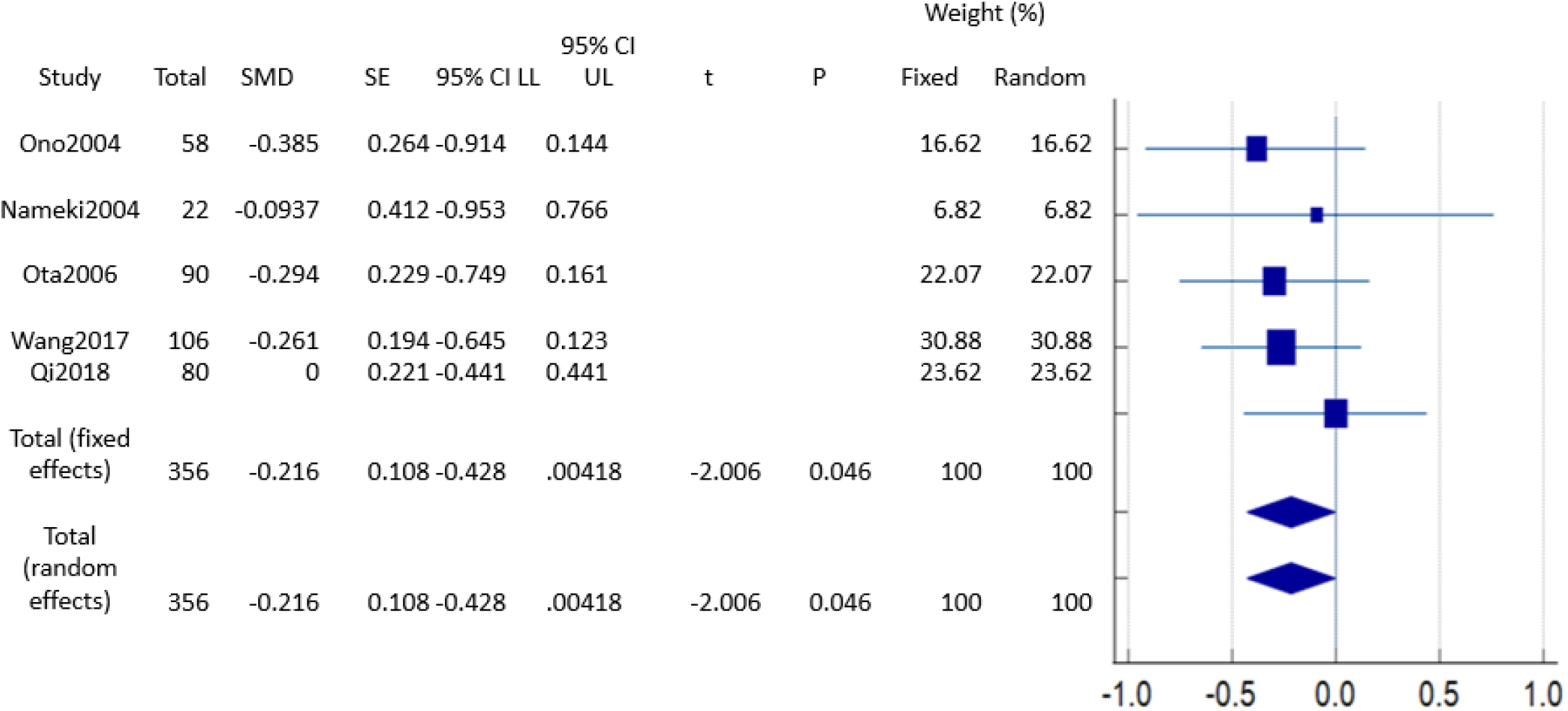
cTFC Comparison 5: - corrected TIMI Frame Count (cTFC)

## Conclusions

With the results it is evident that Nicorandil decreases MACE, NRP while significantly increasing LVEF. It can also be said that it has a protective role in worsening heart failure. All in all, these results suggest that Nicorandil is a highly efficacious drug and should be administered in acute coronary syndrome.

## Data Availability

All data produced in the present work are contained in the manuscript

## List of abbreviations

ACS: Acute Coronary Synderome
PCI: percutaneous coronary intervention
MACE: Major Adverse Cardiac Events
NRP: No-Reflow Phenomenon
LVEF: Left Ventricular Ejection Fraction c
TFC: corrected TIMI Frame Count

---

**Table 1:** Description of papers.

| No. | Author | Type of study | Year of publishing | Place of research | Sample Size | Mean Age | Male/Female | Duration of data collection | Type of cardiac abnormality | Intervention | Quality assessment |
| --- | --- | --- | --- | --- | --- | --- | --- | --- | --- | --- | --- |
| 1 | Hirotsugu Ono(3) | Clinical Trial, RCT | 2004 | Japan | 58 (33 Nicorandil, 25 Control) |  |  |  | Acute Myocardial Infarction | Nicorandil, Control | Good |
| 2 | Mizuo Nameki(12) | Clinical Trial, Comparative Study, RCT | 2004 | Japan | 40 ( Nicorandil, Magnesium, Control) |  |  |  | Acute Myocardial Infarction | Nicorandil, Magnesium, Control | Good |
| 3 | Satoshi Ota(5) | Comparative Study, Multicenter Study, RCT | 2006 | Japan | 92 ( Nicorandil, Nicorandil, Control) |  |  |  | Acute Myocardial Infarction | Intracoronary Nicorandil, Intracoronary and IV Nicorandil, Control | Good |
| 4 | Wang ZQ(14) | RCT | 2017 | China | 158 ( 53 Nicorandil, 52 Nitroglycerin , 53 Control) |  |  | January 2014 to December 2015 | STEMI | Intracoronary Nicorandil, Intracoronary Nitroglycerin, Control | Good |
| 5 | Qi Qi(11) | RCT | 2018 | China | 120 (40 Nicorandil, 40 Sodium Nitroprusside , 40 Control) |  |  |  | Acute STEMI | Intracoronary Nicorandil, Intracoronary sodium nitroprusside , Control) | Good |
| 6 | H Ito(8) | Clinical trial,RCT, comparative study | 1999 | Japan | 81 ( 40 Nicorandil, 41 Control) |  |  |  | Acute Myocardial Infarction | Nicorandil, Control | Good |
| 7 | Tadasu Akagi(19) | RCT | 2006 | Japan | 30 (Nicorandil, Nicorandil, Control) |  |  |  | Acute Myocardial Infarction | IV Nicorandil, IV and Oral Nicorandil, Control | Good |
| 8 | Zuoyuan Chen(9) |  | 2015 | China | 56 |  |  |  | Thirty-six patients with CSF in the left anterior descending (LAD) branch and 20 patients with normal coronary arteries were included. | Nicorandil, Control | Good |
| 9 | Fujiwara (20) | RCT | 2007 | Japan | 62 (31 Nicorandil, 31 Control) |  |  |  | Pre treatment for Myocardial Infarction | Nicorandil, Control | Good |
| 11 | Tsuyoshi Isono(21) | RCT | 2008 | Japan | 49 (Nicorandil, Control) |  |  |  | patients scheduled to undergo elective PCI | Nicorandil, Control | Good |
| 12 | Ju Han Kim(1) | Clinical Trial,RCT | 2005 | Korea | 200 ( 100 Nicorandil, 100 Isosorbide Dinitrate ) | 61 +/- 10 | 143 M , 57 F |  | Unstable Angina | Nicorandil, Isosorbide Dinitrate | Good |
| 13 | Yamada (22) | RCT | 2016 | Japan | 52 |  |  |  | MI patients just before reperfusion | Nicorandil, Control | Good |
| 14 | Akiyoshi Miyazawa( 16) | Eurointervention | 2006 | Japan | 70 ( 35 Nicorandil, 35 control ) |  |  |  | STEMI | Nicorandil, Control | Good |
| 15 | Takuji Toyama(23) | Controlled Clinical Trial | 2006 | Japan | 68 |  |  |  | AMI | Nicorandil, Control | Good |
| 16 | Han Cheon Lee(17) | Clinical Trial, Comparative Study | 2008 | Korea | 73 ( 37 Nicorandil, 36 Control) |  |  |  | Acute STEMI | Nicorandil, Control | Good |
| 17 | Yusuke Kawai(2) | RCT | 2009 | Japan | 408 ( 206 Nicorandil, 202 Control) |  |  |  | ACS and Non ACS patients | Nicorandil, Control | Good |
| 18 | Tory Miyoshi(7) | RCT | 2017 |  | 391 ( Nicorandil, RIPC, Control) |  |  |  | periprocedural myocardial injury | Nicorandil, RIPC, Control | Good |
| 19 | Masaaki Murakami( 4) | RCT | 2006 | Japan | 192 ( 91 Nicorandil, 101 Control) |  |  |  | Patients undergoing coronary stenting | Nicorandil, Control | Good |
| 20 | Young-Guk Ko(13) | Multicenter Study, RCT | 2013 | Korea | 166 ( 81 Nicorandil, 85 Control) |  |  |  | patients with renal dysfunction undergoing coronary angiography . | Nicorandil, Control | Good |
| 21 | Yanming Fan(6) | RCT | 2016 | China | 240 ( 120 Nicorandil, 120 Control) |  |  |  | patients with an estimated glomerular filtration<br>rate (eGFR)<br>of 60<br>mL/min or<br>less, who<br>were<br>undergoing<br>elective<br>cardiac<br>catheterizati<br>on | Nicorandil, Control | Good |

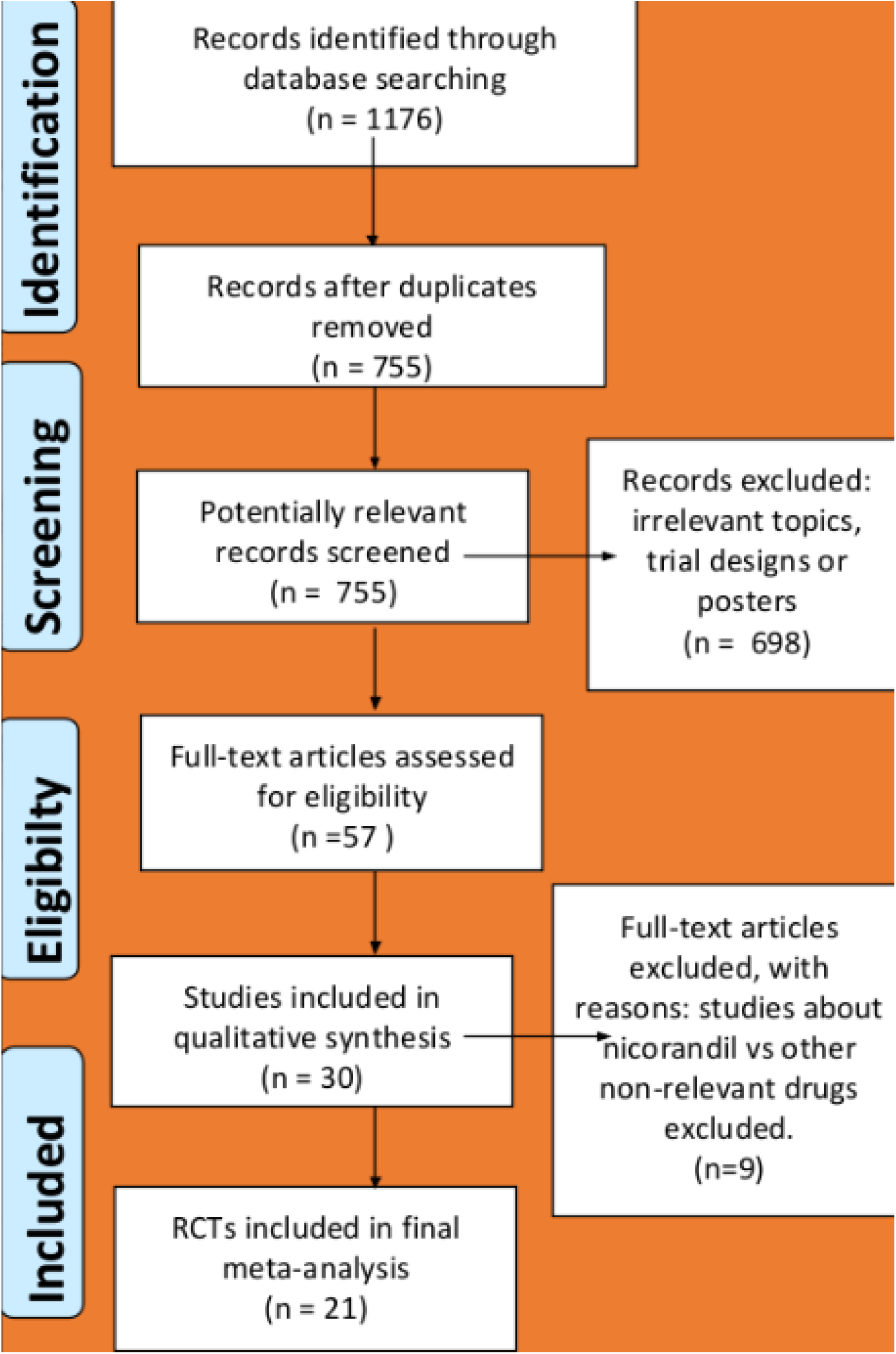
PRISMA Flow Chart

